# Integrated genomic and histopathological analysis of low grade serous ovarian carcinoma identifies clinically distinct disease subtypes

**DOI:** 10.1101/2022.02.01.22270258

**Authors:** John P Thomson, Robert L Hollis, Juliette van Baal, Narthana Ilenkovan, Michael Churchman, Koen van de Vijver, Frederike Dijk, Alison M Meynert, Clare Bartos, Tzyvia Rye, Ian Croy, Patricia Diana, Mignon van Gent, Helen Creedon, Rachel Nirsimloo, Fiona Nussey, Christianne Lok, C. Simon Herrington, Charlie Gourley

**Affiliations:** Nicola Murray Centre for Ovarian Cancer Research, Cancer Research UK Edinburgh Centre, MRC Institute of Genetics and Cancer, University of Edinburgh, Edinburgh, UK; Department of Gynaecologic Oncology and Department of Pathology, The Netherlands Cancer Institute, Antoni van Leeuwenhoek, Amsterdam, The Netherlands; Department of Gynaecologic Oncology and Department of Pathology, Amsterdam University Medical Centres, Amsterdam, The Netherlands; MRC Human Genetics Unit, MRC Institute of Genetics and Cancer, University of Edinburgh, Edinburgh, UK; Edinburgh Cancer Centre, Western General Hospital, Edinburgh, UK

**Author notes:** **Corresponding Author(s):** Professor Charlie Gourley - Nicola Murray Centre for Ovarian Cancer Research, Cancer Research UK Edinburgh Centre, MRC IGC, University of Edinburgh. Crewe Road South, Edinburgh, EH4 2XU, UK. Authors contributed equally (John P Thomson, Robert L Hollis, Juliette van Baal, Christianne Lok, C. Simon Herrington, Charlie Gourley).

## Abstract

**Background:** Low-grade serous ovarian carcinoma (LGSOC) is a distinct, under-investigated and relatively chemotherapy-resistant ovarian cancer type. Understanding the molecular landscape is crucial to maximise the impact of molecularly-targeted therapy.

**Methods:** Whole exome sequencing and copy number data were integrated with histopathological patterns, ER/PR expression, and detailed clinical annotation, including survival, in a carefully curated LGSOC cohort.

**Results:** 63 tumours were analysed in the largest comprehensive genomic LGSOC study to date. Three genomic subgroups were identified: canonical MAPK mutant (cMAPKm: 52%, *KRAS*/*BRAF*/*NRAS*), MAPK-associated mutation (27%, 14 MAPK-associated genes) and MAPK wild-type (MAPKwt: 21%). MAPKwt patients were younger at diagnosis (median 47 versus 62 years in the cMAPKm subgroup) and demonstrated shorter survival [multivariable HR (mHR) 4.17]. The inferior survival in the MAPKwt subgroup was due to shorter post-relapse survival (mHR 5.22) rather than shorter time to first progression (mHR 1.15). Patients in the MAPK-associated mutation subgroup had similar survival to cMAPKm cases. The cMAPKm subgroup more frequently demonstrated macropapillary invasion. Desmoplasia and micropapillary invasion were independently associated with poor survival. NOTCH pathway activation occurred independently of MAPK subgroup.

**Conclusions:** LGSOC comprises multiple genomic subgroups with distinct clinical, molecular and histopathological features. True MAPKwt cases represent around a fifth of patients: they are younger but have poorer survival. New therapeutic strategies with activity in this subgroup are urgently required. NOTCH inhibitors represent a therapeutic strategy worthy of exploration.

## Introduction

Low grade serous ovarian carcinoma (LGSOC) is an uncommon (<5%) and under-investigated ovarian cancer type, in some cases arising from serous borderline tumours [1]. In contrast to high grade serous ovarian cancers (HGSOCs), LGSOCs are *TP53* wild-type, do not demonstrate genomic instability or homologous recombination repair deficiency and frequently harbour classical activating mutations in canonical MAPK pathway components (*KRAS, BRAF* and *NRAS*) [2-5]. LGSOCs typically display high oestrogen receptor (ER) expression, with expression of progesterone receptor (PR) in a subset of cases [6].

LGSOC often affects younger women (median 47 years) [7]. The response rate to platinum-based chemotherapy is 5-25% [8, 9], compared to 70-80% in HGSOCs. Relapsed LGSOC follows a more indolent course than relapsed HGSOC, demonstrating prolonged post-relapse survival [10, 11]. Appreciation of LGSOC as a separate entity has driven a management shift away from the conventional ‘one size fits all’ approach to ovarian cancer treatment, with endocrine therapy showing significant activity [12, 13]. Recently, the MEK inhibitor trametinib also demonstrated notable efficacy, delaying disease progression in the context of LGSOC recurrence compared to standard of care therapies [14, 15]. Another MEK inhibitor binimetinib, while not demonstrating superiority to standard of care chemotherapy, did show an overall response rate of 16% with a suggestion that patients whose tumours harboured activating *KRAS* mutations were more likely to respond [16].

Until recently, genomic characterisation of LGSOC had been limited to targeted sequencing of selected genes [3, 4, 17-19] or whole exome sequencing of a small number of cases (n=8, n=9, n=22) [5, 17, 20]. Most recently, a targeted sequencing study of 71 LGSOC cases [19] identified mutation of *KRAS, BRAF* or *NRAS* in 47% of cases, alongside further less common potentially actionable mutations.

Here we investigate whether comprehensive genomic characterisation of LGSOC can be implemented to stratify LGSOC cases into clinically meaningful subgroups. We perform whole exome sequencing (WES) on tumour material from 63 LGSOC patients, identifying genomic subgroups centred around the MAPK pathway. These subgroups demonstrate significant differences in patient age, clinical outcome and histopathological features; we identify patient groups more likely to experience long-term survival, and highlight high-risk patient groups for which new treatment strategies are required to improve outcome.

## Methods

### Ethical approval

Ethical approval was obtained from the Lothian Human Annotated Bioresource (reference 15/ES/0094-SR925), NKI-AVL Translational Research Board (reference CFMPB284), and University of Amsterdam AMC Biobank Assessment Committee (reference 2016_070#A201641). All participants provided written informed consent or had consent waived by the ethics committee due to the retrospective nature of the study.

### Cohort identification and pathology review

Cases were identified from local databases. All cases underwent expert gynaecological pathology review (KvdV, CSH). Cases were excluded if: WT1 negative; aberrant p53 expression; no definitive stromal invasion; *TP53* gene mutation.

### Assessment of histopathological patterns in LGSOC

H&E-stained sections underwent secondary review to identify the presence of desmoplasia and the macropapillary (MaP) and micropapillary (MiP) patterns of invasion[22]. MaP was defined as stromal invasion by papillary structures containing fibrovascular cores[22]; MiP was defined as invasion by papillary structures without fibrovascular cores.

### Clinical data

Baseline patient characteristics, treatment information and outcome data were retrieved from local databases and from patient file review. Disease-specific survival (DSS) was calculated from the date of confirmed ovarian cancer (OC) diagnosis.

### Whole exome sequencing, variant calling and analysis

Following DNA extraction from macrodissected FFPE sections (Supplementary Methods 1), exome capture was performed using the Illumina TruSeq Exome Library Prep kit and WES was performed on the Illumina NextSeq 550 (Illumina, Inc., San Diego, CA, USA). Samples failed sequencing QC if mean on-target coverage was <30X. Data were aligned to the GRCh38 human reference genome using bwa-0.7.17[29], duplicates marked and base quality scores recalibrated with the GenomeAnalysisToolkit (GATK) v4[30] within the bcbio 1.0.6 pipeline (Supplementary Methods 2). Variants were called using a majority vote system from three callers (VarDict[31], Mutect2[32] and Freebayes[33]) and were filtered to remove common and likely non-functional variation. Analysis of variants and oncogenic pathways was performed using the R package maftools[34]. Functional tumour mutational burden (fTMB) was calculated as the number of mutations present following filtering. CNV analysis was carried out using a custom script utilising the Python package multiBamSummary, comparing read normalised 10kb binned scores between in house “normal” buffy coat derived sample datasets and LGSOC samples. For more information on these analyses see Supplementary Methods 3-6.

### Immunohistochemistry for ER and PR

Immunohistochemistry for ER and PR was performed using protocol F on the Leica BOND III Autostainer (Supplementary Methods 7). ER immunohistochemistry used rabbit anti-ER antibody M3643 clone EP1; PR immunohistochemistry used mouse anti-PR antibody M3569 clone PgR-636. Nuclear expression was quantified using histoscore, generated by multiplying the proportion of positive tumour nuclei (0 – 100%) by the intensity of nuclear staining (0 – 3) to produce weighted scores from 0 to 300[35]. Two independent observers scored digital images of stained slides, demonstrating excellent agreement (rho=0.96 for PR, rho=0.93 for ER) (Supplementary Methods 7). Final patient histoscore was calculated as the mean score of the two observers.

### Statistical analysis

Statistical analyses were performed using R version 4.0.3. The Mann Whitney-U test or T-test were used to compare continuous data, as appropriate. Survival analyses were performed using the Cox proportional hazards regression analysis in the Survival package. Univariable survival analyses are presented as hazard ratios (HRs) alongside 95% confidence intervals. Multivariable analyses are presented similarly, detailing multivariable hazard ratios (mHRs). Median follow-up time was calculated using the reverse Kaplan-Meier method. Comparisons of frequency were performed using the Chi-squared test or Fisher’s exact test, as appropriate.

### Data availability

The genomic data presented in this manuscript are being made available via deposition in the European Genome-Phenome Archive.

## Results

### Cohort characteristics

118 LGSOC cases were identified. 41 cases were excluded for insufficient tumour material or cellularity; 3 failed DNA quality control; 5 had insufficient sequencing coverage. 69 cases underwent genomic characterisation by WES. 6 cases harbouring *TP53* mutations were excluded as potential occult HGSOC, leaving 63 cases in the final LGSOC study cohort (Figure S1). The median per-sample on-target coverage in the study cohort was 64X (range 32X-132X).

Clinical characteristics of the LGSOC study cohort are summarized in Table 1. The median follow-up time was 13.3 years. All patients underwent cytoreductive surgery. The median disease-specific survival (DSS) was 12.9 years.

**Table 1.**
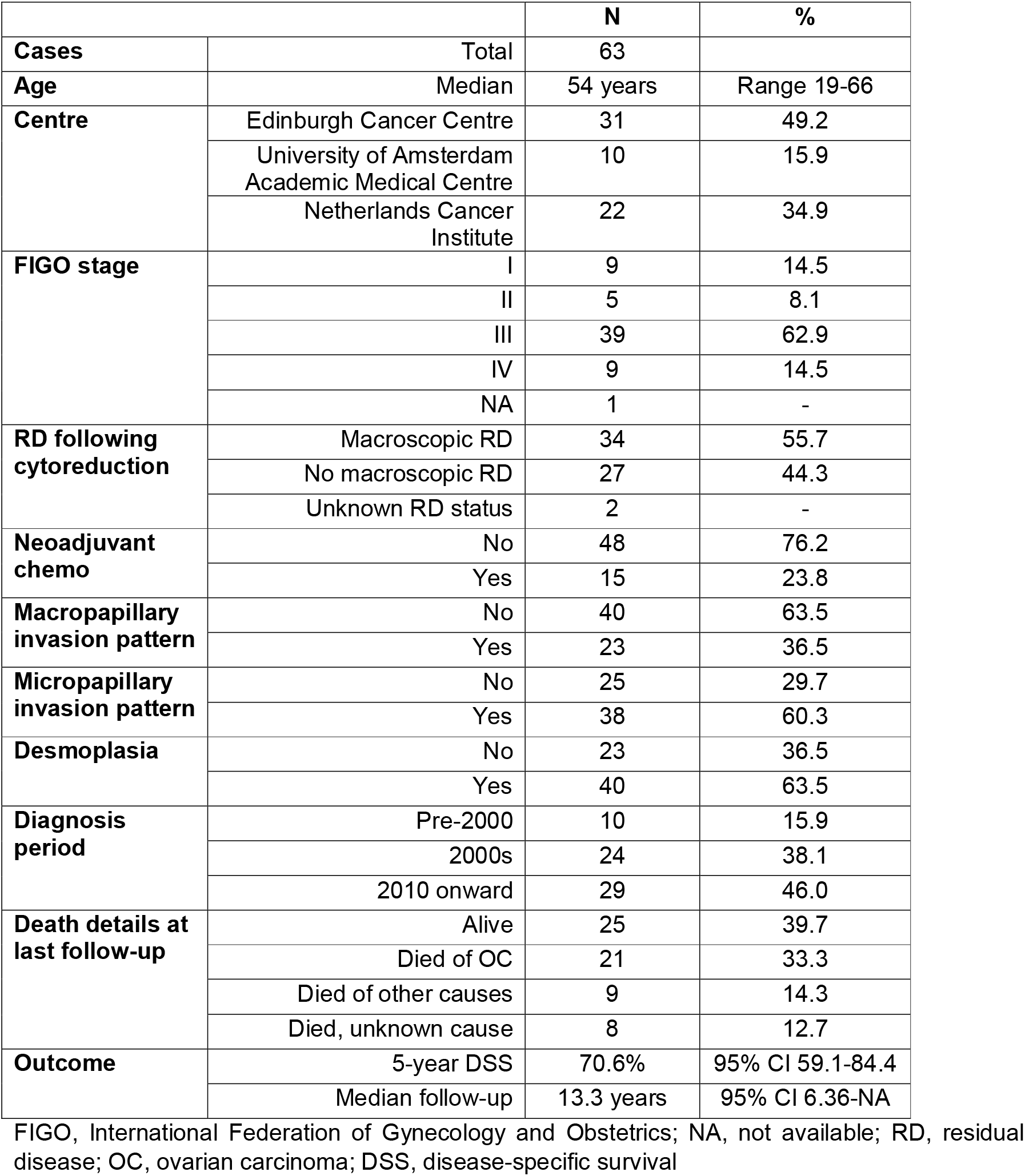
Clinical characteristics of low grade serous ovarian carcinoma cases.

### Genomic features identify subgroups of low grade serous ovarian carcinoma

WES identified canonical MAPK pathway mutations in 33 cases (52.4%) (cMAPKm subgroup): 24 *KRAS* (38.1%), 6 *BRAF* (9.5%) and 3 *NRAS* (4.8%) (Figures 1, S2, S3), which occurred mutually exclusively. The majority of these events occurred at known mutational hotspots (83% of *KRAS* G12R/C/D/V, 100% *BRAF* V600E, 100% *NRAS* Q61R/K) (Figure S4).

**Figure 1:**
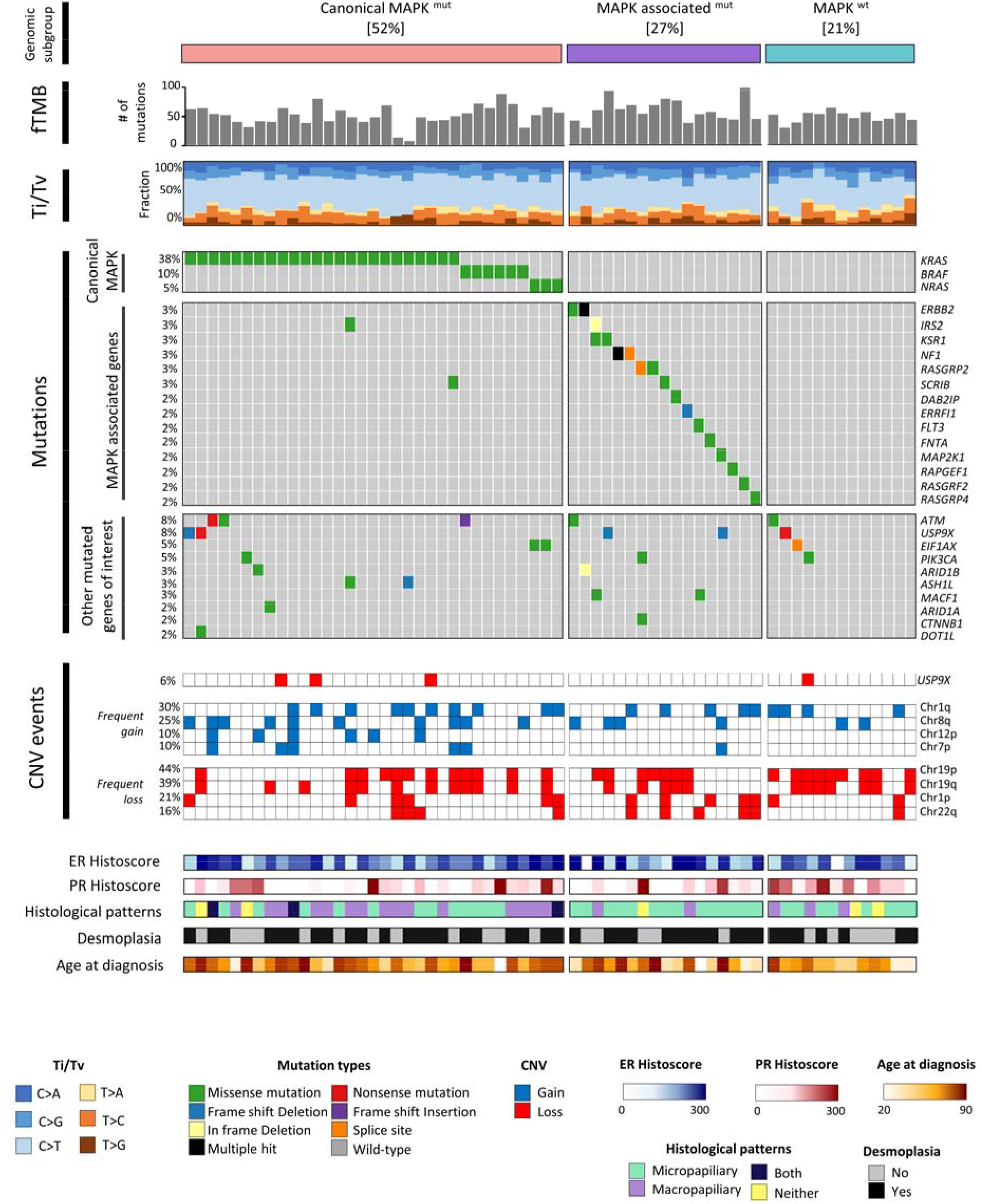
Molecular landscape of low grade serous ovarian carcinoma. fTMB, functional tumour mutational burden; Ti, transitions; Tv, transversions; SNV, single nucleotide variant; CNV, copy number variant ; cMAPKm, canonical MAPK pathway mutation (KRAS/BRAF/NRAS); MAPK-assoc, MAPK-associated mutation; MAPKwt, MAPK pathway wild-type.

Pathway analysis of mutated genes revealed a further 17 cases (27.0%) with perturbation of the MAPK/RAS pathway by mutations across 14 other MAPK-associated genes defined by the TCGA PanCanAtlas [21] (MAPK-associated mutations) (Figures 1 S5). These events were largely mutually exclusive (co-occurrence in 1 case), and genomic events in these genes seldom occurred in cMAPKm cases (co-occurrence in 2 cases). The total frequency of MAPK/RAS pathway alteration was 50 of 63 cases (79.4%) (33 cMAPKm, 17 MAPK-associated mutations).

A further 13 cases were MAPK wild-type (MAPKwt). These patients demonstrated significantly younger age at diagnosis compared to cMAPKm cases (median 47 vs 62 years, P=0.048) (Figures 1, 2A & Table S1).

**Figure 2.**
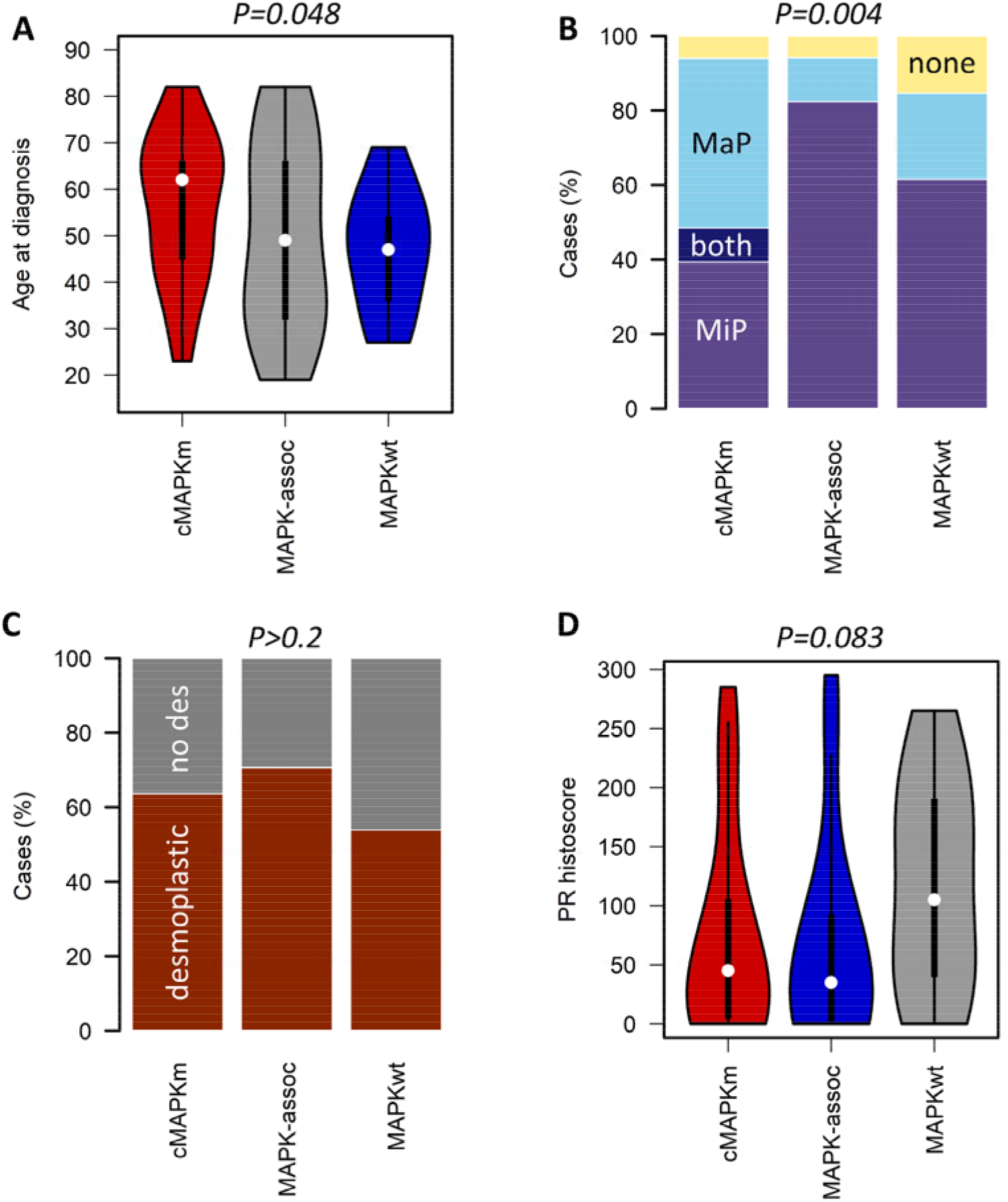
Characteristics of genomic subgroups in low grade serous ovarian carcinoma. (A) Patient age at diagnosis across genomic subgroups; labelled P value represents the comparison between the canonical MAPK mutation (cMAPKm) group and the MAPK wild-type (MAPKwt) group. (B) Histopathological patterns across genomic subgroups; labelled P value represents comparison of MaP frequency in cMAPKm versus subgroups by Chi-squared test. (C) Desmoplasia across genomic subgroups; labelled P value represents comparison of desmoplasia frequency between all subgroups by Chi-squared test. (D) Progesterone receptor (PR) expression across genomic subgroups; labelled P value represents comparison of MAPKwt group versus other subgroups by Mann Whitney-U test. MiP: micropapillary; MaP: macropapillary. No des = no desmoplasia identified.

NOTCH pathway perturbation was also common (21.0% of cases), occurring both in the context of MAPK/RAS pathway alteration (34.0%, 17/50) and in MAPKwt cases (30.8%, 4/13) (Figure S5). Recurrent gene targets of mutation beyond MAPK pathway components included *ATM* (5 cases, 7.9%), *USP9X* (5 cases, 7.9%) and *EIF1AX* (3 cases, 4.8%) (Figures 1, S3). There was significant co-occurrence between *NRAS* and *EIF1AX* mutation (P=0.005).

### Copy number alterations in low grade serous carcinoma

Across the cohort, loss of all or part of chromosome 19 (chr19) was common (22 cases (34.9%): p and q arms; 6 cases (9.5%): p arm; 2 cases (3.2%): q arm). Chr19p loss was most frequent in MAPKwt cases (9 of 13 cases, 69.2%), but this was not statistically significantly higher than in the cMAPKm group (P=0.056) (Figure S6).

Gain of the q arm of chr1 occurred in 19 cases (30.2%), while loss of the p arm of chr1 occurred in 13 cases (20.6%). Gain of the q arm of chr8 was also common (16 cases, 25.4%). In agreement with previous reports, four cases (6.3%) demonstrated copy number loss specifically over the *USP9X* locus.

### Histopathological patterns in low grade serous carcinoma

A MiP pattern of invasion was identified in 38 cases (60.3%), while a MaP invasion pattern was identified in 23 cases (36.5%) (Figure 1)[22]. The MiP and MaP patterns were largely mutually exclusive (P<0.0001, co-occurrence in only 3 cases). 5 cases demonstrated neither MaP nor MiP pattern. The MaP pattern was more common in cMAPKm cases (54.5%) compared to MAPKwt (23.1%; p=0.004; Figure 2B).

Desmoplasia was identified in 40 cases (63.5%). Desmoplasia was associated with advanced stage at diagnosis (92.5%, 37 of 40 desmoplastic cases with FIGO III/IV vs 50.0%, 11 of 22 evaluable non-desmoplastic cases with FIGO III/IV; P<0.001). Desmoplasia was also associated with the presence of macroscopic RD (67.5%, 27 of 40 desmoplastic cases demonstrated macroscopic RD vs 33.3%, 14 of 21 evaluable non-desmoplastic cases, P=0.015) (Figures 2C, 3A, 3B).

**Figure 3.**
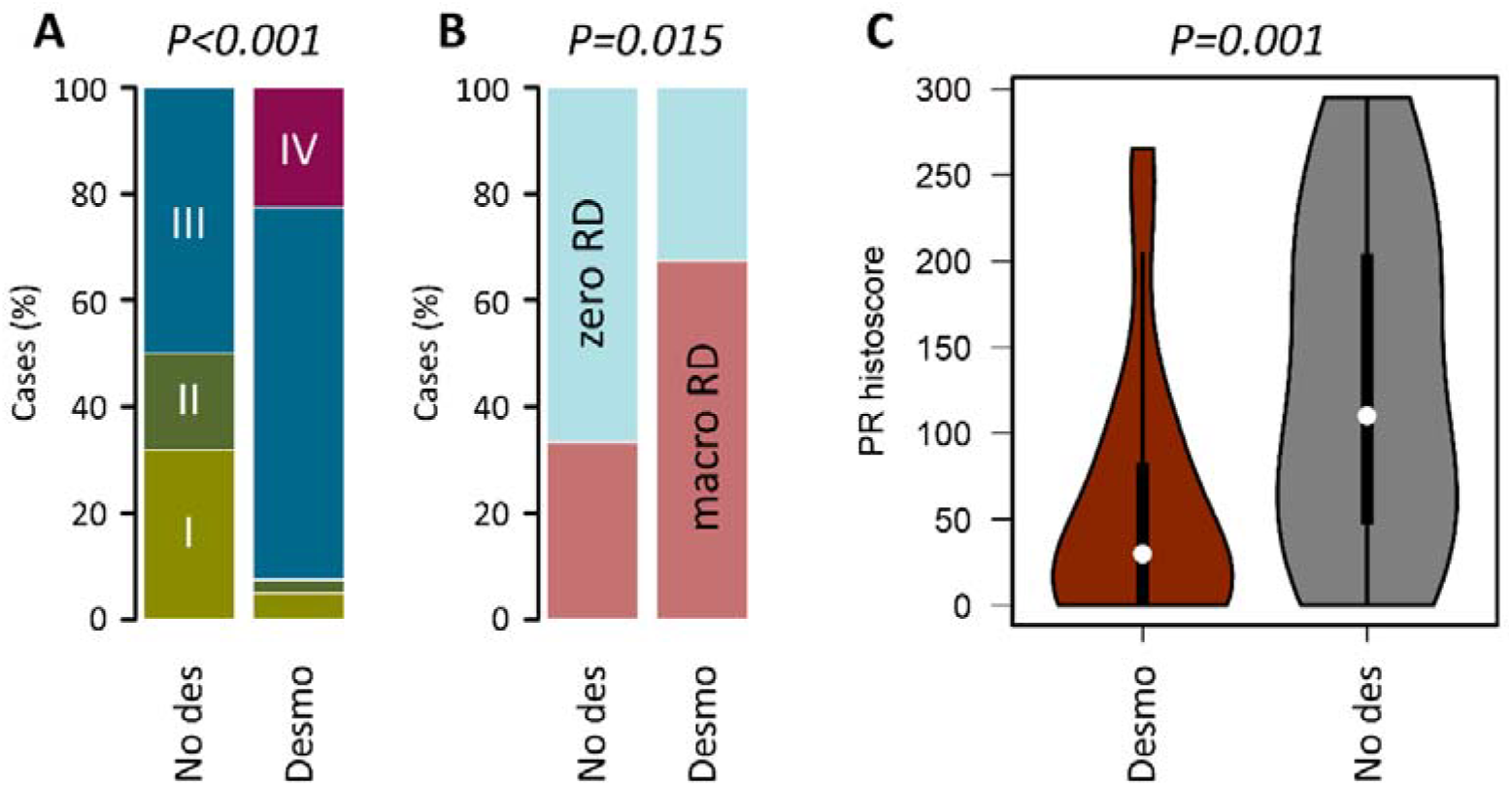
Features associated with desmoplasia in low grade serous ovarian carcinoma. (A) Distribution of stage at diagnosis between tumours with and without desmoplasia. (B) Distribution of stage at diagnosis between tumours with and without desmoplasia. (C) Progesterone receptor (PR) expression between cases with and without desmoplasia. Desmo = desmoplastic; No des = no desmoplasia identified; zero RD = zero macroscopic residual disease after debulking surgery; macro RD = macroscopic residual disease after debulking surgery.

### ER and PR expression in low grade serous carcinoma

LGSOC samples stained uniformly for ER (histoscore ≥50 in 60 of 61 evaluable cases, 98.4%; 2 non-evaluable) (Figure S7A). Median ER and PR histoscores were 220 (range 7.5-300) and 50 (range 0-295) respectively (Figure S7A and S7B). Analysis of ER and PR levels across the genomic classes revealed higher levels of PR expression in the MAPKwt subgroup (median PR histoscore 105) (Figure 2D, Figure S7C), but this was not significantly higher compared to the cMAPKm and MAPK-associated subgroups (median PR histoscore 45 and 35, P=0.083).

Desmoplastic tumours demonstrated significantly lower PR expression (median histoscore 30 vs 110, Bonferroni-adjusted P=0.003) (Figure 3C) than non-desmoplastic tumours.

### Features associated with outcome in low grade serous carcinoma

Patients with LGSOC demonstrating desmoplasia experienced significantly shorter DSS (HR for no desmoplasia 0.18, 95% CI 0.05-0.62) (Figures 4A, S8A); the MiP pattern was also associated with poor DSS (HR for absence of MiP 0.29, 95% CI 0.10-0.86) (Figures 4B, S8B). Conversely, high PR expression was associated with significantly prolonged survival (HR for PR histoscore ≥50 0.23, 95% CI 0.09-0.60) (Figures 4C, S8C).

**Figure 4.**
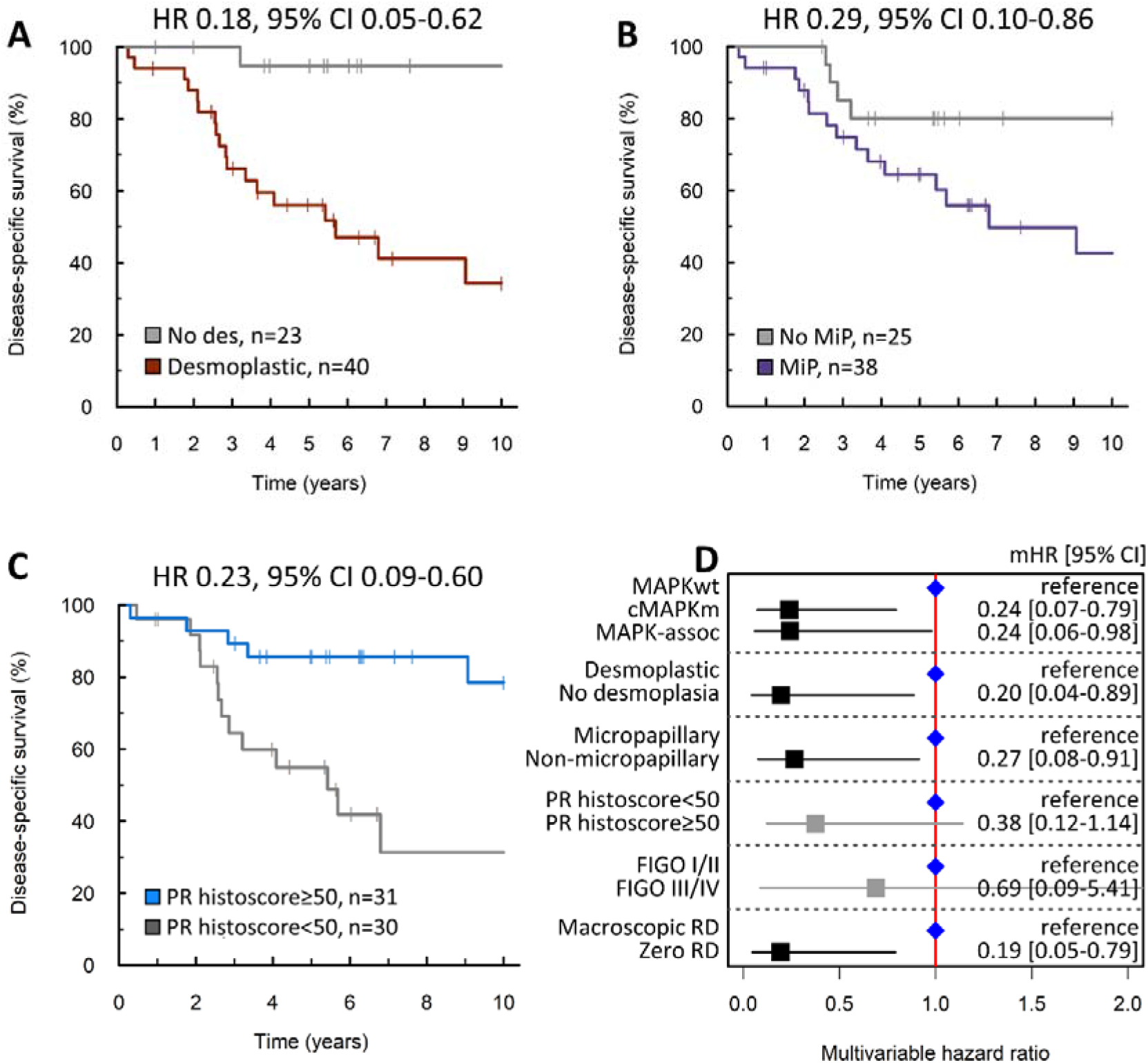
Impact of low grade serous ovarian carcinoma features on disease-specific survival. (A) Disease-specific survival (DSS) of patients with tumours demonstrating desmoplasia. (B) DSS of patients with tumours demonstrating the micropapillary (MiP) pattern of invasion. (C) DSS of patients with tumours expressing progesterone receptor (PR). (D) Forest plot summarising multivariable survival analysis of low grade serous ovarian carcinoma features. HR, hazard ratio; 95% CI, 95% confidence interval; mHR, multivariable HR; no des, no desmoplasia identified; RD, residual disease; cMAPKm, canonical MAPK pathway mutation (KRAS/BRAF/NRAS); MAPK-assoc, MAPK-associated mutation; MAPKwt, MAPK pathway wild-type.

Multivariable analysis of LGSOC features (genomic subgroup, PR status, MiP pattern and desmoplasia) accounting for clinicopathological variables identified the MAPKwt subgroup (multivariable HR [mHR] for cMAPKm vs MAPKwt 0.24, 95% CI 0.07-0.79), desmoplastic cases (mHR for absence of desmoplasia 0.20, 95% CI 0.04-0.89) and the MiP pattern (mHR for absence of MiP=0.27, 95% CI 0.08-0.91) as independently associated with shorter DSS (Figure 4D, Table S2). PR status was not independently associated with significantly longer DSS (mHR 0.38, 95% CI 0.12-1.14).

Further endpoint analysis suggested that inferior DSS in the MAPKwt group was due to shorter post-relapse survival (mHR 5.22, 95% CI 1.20-22.67) rather than rapid disease progression following first-line treatment (mHR 1.15, 95% CI 0.43-3.08).

## Discussion

LGSOC affects younger women than HGSOC and demonstrates greater intrinsic chemoresistance. Although survival is better than HGSOC, patients frequently relapse resulting in morbidity from largely ineffectual therapies and premature mortality. As such, better (molecular) characterization and new therapies are urgently required.

Few studies have performed molecular characterisation of robustly curated LGSOC cohorts. Where molecular profiling studies have been performed, these have been limited to panel based sequencing of selected genes [3, 18, 19], or have included only a small number of patient samples [5, 20]. Few of these studies have used immunohistochemistry for WT1/p53 to confirm LGSOC diagnosis. Here we report comprehensive genomic profiling by WES, integrating these data with histopathological analysis, clinical data and assessment of ER and PR expression patterns in a sizeable LGSOC cohort.

52% of cases harboured mutations in the canonical MAPK pathway components *KRAS, BRAF* and *NRAS*. This cMAPKm group more frequently demonstrated the MaP pattern of invasion, represented older patients (median 62 years) and was associated with more favourable DSS and post-relapse survival. These data are consistent with previously reported favourable survival in LGSOC patients with tumours harbouring *KRAS* or *BRAF* mutations identified by targeted sequencing [3]. Our data suggest that this survival benefit is due to prolonged post-relapse survival.

While the rate of cMAPKm within our LGSOC cohort (52%) is similar to that reported in previous targeted sequencing studies [17, 19] and WES of small LGSOC cohorts [20], we also identified a further 27% of cases (MAPK-associated group) demonstrating mutational disruption of genes related to the MAPK pathway [21]; these cases demonstrated a survival profile similar to that of the cMAPKm cases. The remaining MAPKwt cases (21%) represented patients with younger age at diagnosis (median 47 years) and who experienced shorter survival. This suggests that previous observations of poorer survival in younger LGSOC patients may be underpinned by differences in disease biology between younger and older patients [23]. MAPKwt cases often demonstrated the MiP pattern of invasion (62%), with only 23% demonstrating the MaP pattern.

While different histopathological patterns of invasion (MaP and MiP) were associated with the genomic subgroups, we demonstrate that the MiP pattern is independently associated with shorter survival time (mHR 3.76). We also demonstrate that the presence of desmoplasia is associated with poorer outcome (mHR 5.10), independent of its relationship with more advanced stage disease and residual disease status.

Both the GOG281/LOGS and MILO studies suggested that the response rates to MEK inhibition were greater in patients with canonical MAPK defects [14, 16]. However, to date, no assessment of relative MEK inhibitor efficacy has been made with regard to MAPK pathway mutations beyond *KRAS*/*BRAF*/*NRAS*; future work should seek to quantify efficacy within these patients, and should determine whether patients with no identifiable genomic MAPK defect (MAPKwt) derive any benefit from MEK inhibition.

Beyond the MAPK pathway, analysis of genomic events from our WES data suggest perturbation of the NOTCH signalling pathway is common, affecting a notable proportion of cases both with and without MAPK defects (34% and 31%, respectively). NOTCH pathway inhibition may therefore represent a novel therapeutic strategy for LGSOC patients with intrinsic or acquired resistance to chemotherapy and/or MEK inhibitors.

We report the first investigation using WES in a sizeable LGSOC cohort (n=63) with detailed clinical annotation and extensive follow-up. The use of immunohistochemistry for WT1 and p53 status to robustly curate our cohort, represents a major strength of this work. However, while our dataset represents one of the largest LGSOC collections reported to date, the overall cohort size remains modest due to the rarity of LGSOC. Confirmatory studies characterising the behaviour of the described genomic subgroups, and associations with age, histopathological features and outcome, are therefore required.

Collectively, our data identify a subgroup of LGSOC with mutations in MAPK pathway components beyond *KRAS, BRAF* and *NRAS*. Alongside the cMAPKm subgroup, these cases demonstrate more favourable outcome when compared to MAPKwt patients, who are younger at diagnosis. The MaP pattern of invasion is a potential histopathological indicator of cMAPKm LGSOC. Desmoplasia and the MiP pattern are features independently associated with survival.

## Data Availability

The genomic data presented in this manuscript are being made available via deposition in the European Genome-Phenome Archive.
We are happy to provide data to researchers upon reasonable request and in compliance with our research ethics framework.

## Acknowledgements

Funding: RLH was supported by Target Ovarian Cancer. This work was funded in part by a HANARTH Foundation grant. We extend our thanks to The Nicola Murray Foundation for their generous support of The Nicola Murray Centre for Ovarian Cancer Research. JT and CB were supported by core funding from the CRUK Edinburgh Centre. AMM was supported by core funding awarded to the MRC Human Genetics Unit.

We are grateful to the NHS Lothian Department of Pathology, Edinburgh Experimental Cancer Medicine Centre, the Edinburgh Ovarian Cancer Database and the NRS Lothian Human Annotated Bioresource for their ongoing support. The high throughput sequencing described here was supported by the Edinburgh Clinical Research Facility, Western General Hospital, Edinburgh, UK. We would like to acknowledge the NKI-AVL Core Facility Molecular Pathology & Biobanking (CFMPB) for supplying NKI-AVL Biobank material and providing laboratory support.

