## supplementary materials for "Integrated genomic and histopathological analysis of low grade serous ovarian carcinoma identifies clinically distinct disease subtypes"

**Supplementary Materials and Methods**

1. ***DNA extraction***

Nucleic acid extraction was performed using 10µm FFPE sections, macrodissected to enrich for tumour purity. Macrodissection was guided by H&E stained slides marked by an expert gynaecological pathologist (CSH). DNA extraction was performed using the QIAamp DNA FFPE Tissue Kit (Qiagen) and Deparaffinization Solution according to the manufacturer’s instructions.

2. **Mapping of sequenced reads**

Base calling and quality were assessed using FASTQC. Data were processed with the bcbio-nextgen python toolkit for fully automated high throughput sequencing analysis (see <https://github.com/bcbio/bcbio-nextgen>). Raw sequence data was mapped to the hg38 genome build using the Burrows–Wheeler alignment algorithm 0.7.17 (1).

3. ***Variant calling and classification***

Following alignment to the hg38 reference genome, variant calling was performed using a majority vote system from three variant caller algorithms: VarDict(2), Mutect2(3) and Freebayes(4). Filtering for C>T (FFPE artifacts) and G>T (oxidation artifacts) was applied using GATK (CollectSequencingArtifactMetrics and FilterByOrientationBias). Variants associated with low sequence depth (<20X) or low variant allele frequency (<10%) were removed. Common variants were excluded using the 1000 genomes and ExAC reference datasets; known pathogenic and benign variants were flagged using ClinVar (5), and remaining variants were filtered to remove likely non-functional variation using the Polymorphism Phenotyping (PolyPhen) (6) and Sorting Intolerant from Tolerant (SIFT)(7) functional prediction tools.

4. ***Analysis of SNVs***

Variants were visualised using the R package maftools (8). Transitions and transversions were calculated using the *Titv* function.

5. ***Pathway analysis***

Pathway analysis was carried out using the OncogenicPathways function in maftool (8), comparing input variants against oncogenic signalling pathway gene lists defined by Pan-cancer TCGA analysis (9). Functional analysis was carried out through the use GOTERM_BP_FAT function in The Database for Annotation, Visualization and Integrated Discovery (DAVID) at <https://david.ncifcrf.gov/home.jsp> (10). Functional terms were filtered to remove values where the value >0.05 against expected values and then ranked by the proportion of genes in a given term. Plots were selected to show at least the top 15 terms with gene set proportions > 0.5%.

6. ***Analysis of CNVs***

A custom script was run to derive copy number variation (CNV) events in the absence of matched normal samples. In brief, sliding window analysis was carried out on aligned BAM files using the Python package multiBamSummary (python v3.7.3, multiBamSummary via deepTools v2.0), using a 10kb bin size. A normal background dataset was generated using in house buffy coat sequencing data from 3 individuals. Both “normal” and LGSOC samples were then normalised for total mapped read count and then binned LGSOC signal values subtracted from the similarly binned normal background signal. To derive focal CNVs the resulting binned values were mapped to hg38 refgene gene coordinates and average binned signals across a gene calculated. Focal CNV changes were defined as genes which deviated by greater than 3 standard deviations from the cohort average gene values. To derive regional CNV changes 10kb binned datasets were taken and the average value across entire chromosome arms calculated using UCSC HG38 cytoband annotations. Regional CNV changes were defined as arms in which the average signal was at least 1.5 fold greater than or less than the normal backbone signal. For the purposes of this study and based on the findings of previous studies (11), we restricted our analysis to only consider CNV events over the *USP9X* locus as well as regional CNV changes at the chromosomal arm level.

7. ***Immunohistochemistry for ER and PR***

Immunohistochemistry for ER and PR was performed using protocol F on the Leica BOND III Autostainer using epitope retrieval solution 2 for 20 minutes. ER IHC used rabbit anti-ER antibody M3643 clone EP1; PR IHC used mouse anti-PR antibody M3569 clone PgR-636. Normal human breast tissue was used as a positive control for both markers. Nuclear ER and PR expression was assessed by histoscore, a quantitative nuclear expression score generated by multiplying the proportion of positive tumour nuclei (0 – 100%) by the intensity of nuclear staining (0 – 3) to give weighted scores from 0 to 300 (12). Histoscoring by two independent observers (CSH, RLH) demonstrated excellent agreement (ρ=0.96 for PR, ρ=0.93 for ER), with a median histoscore difference of 10 and 20 for PR and ER, respectively. Final patient histoscore was calculated as the mean score of the two observers.

**Supplementary Figures**

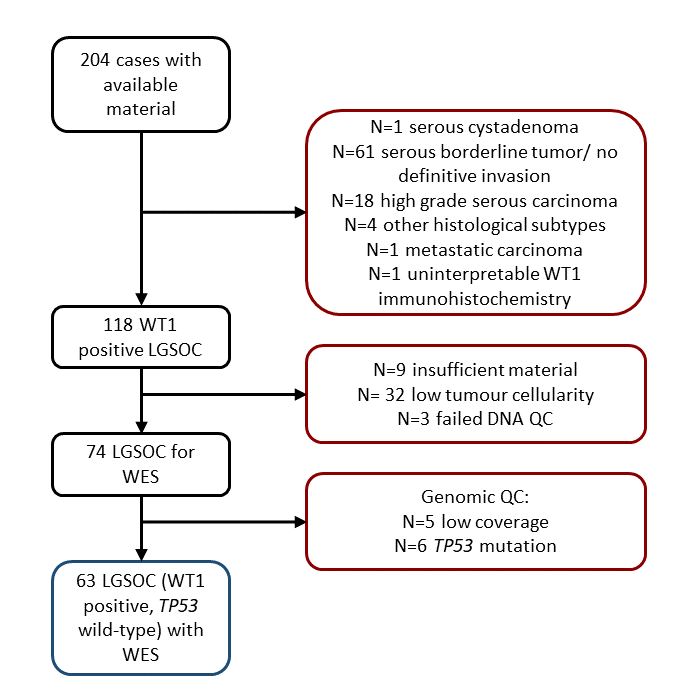

Figure S1. Case flow diagram identifying 63 low grade serous ovarian carcinomas characterized by whole exome sequencing. LGSOC, low grade serous ovarian carcinoma. WES, whole exome sequencing. QC, quality control.

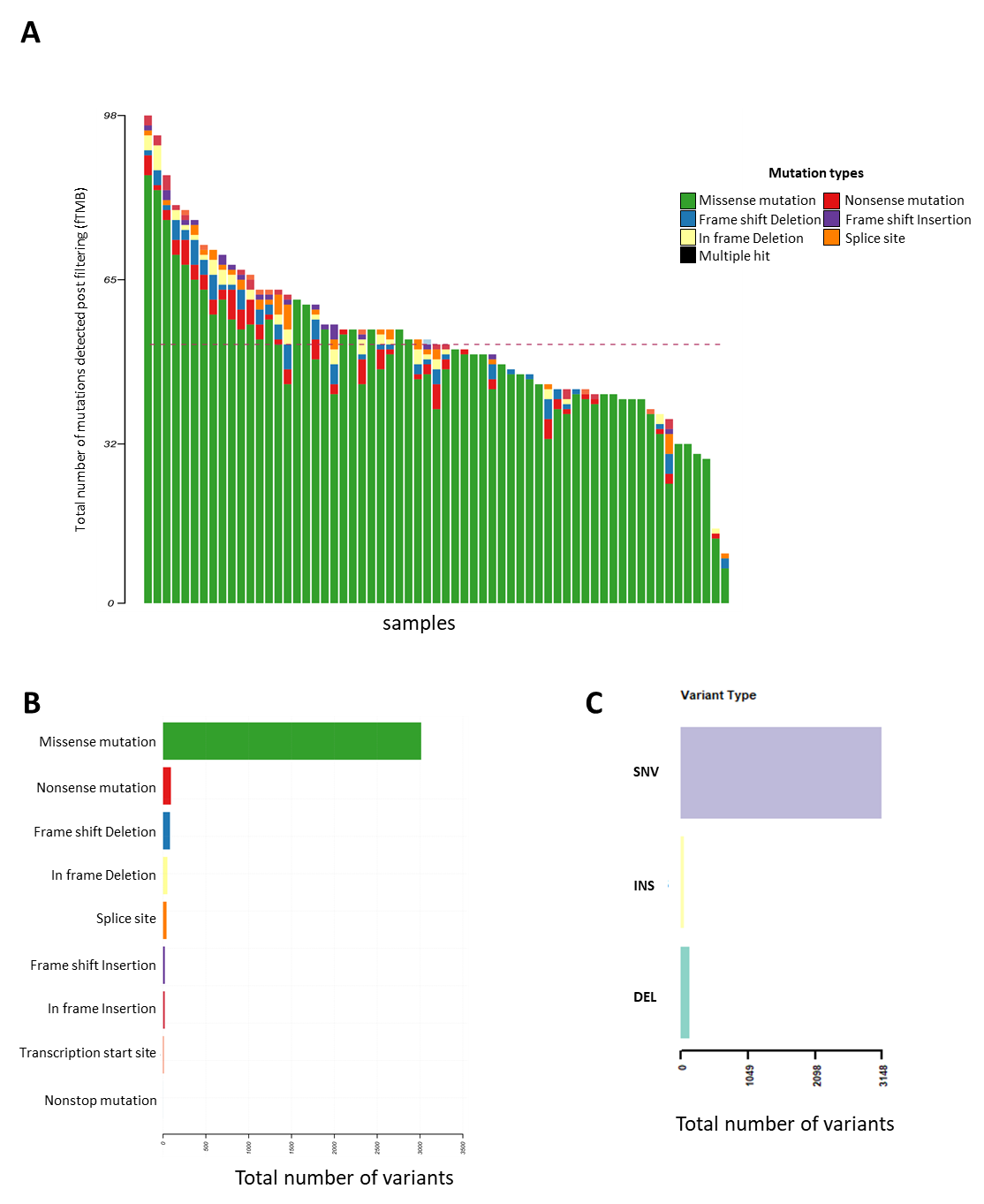

Figure S2. Whole exome variant call summary statistics. A. Total number of variants detected across samples after filtering (functional tumour mutational burden: fTMB). B. Summary of single nucleotide variant (SNV) types. C. Total SNV, insertions (INS) and deletions (DEL) across all samples.

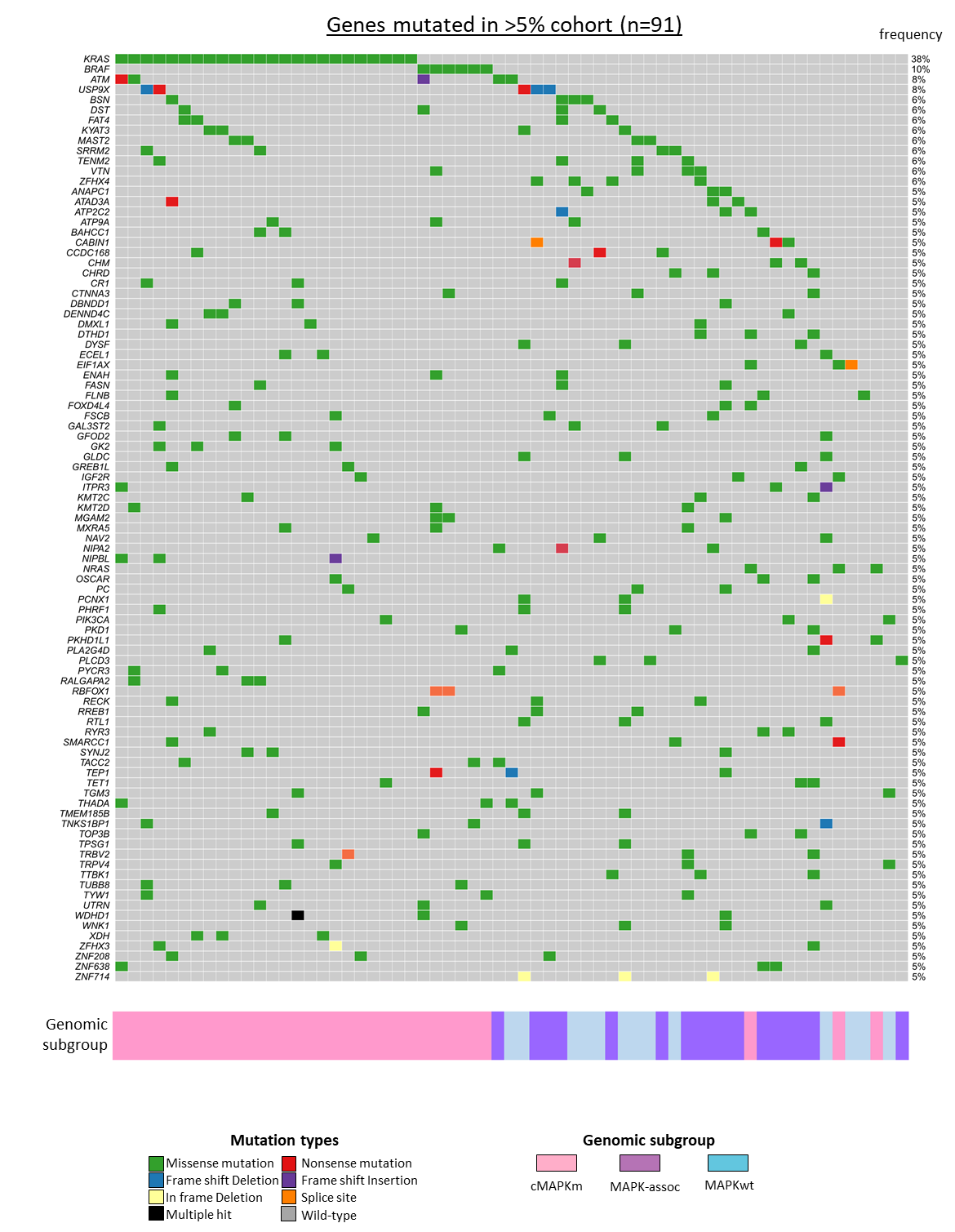

Figure S3. Oncoplot of mutated genes found mutated in at least 5% of samples. Grey denotes wild-type. Genomic subgroup shown below.

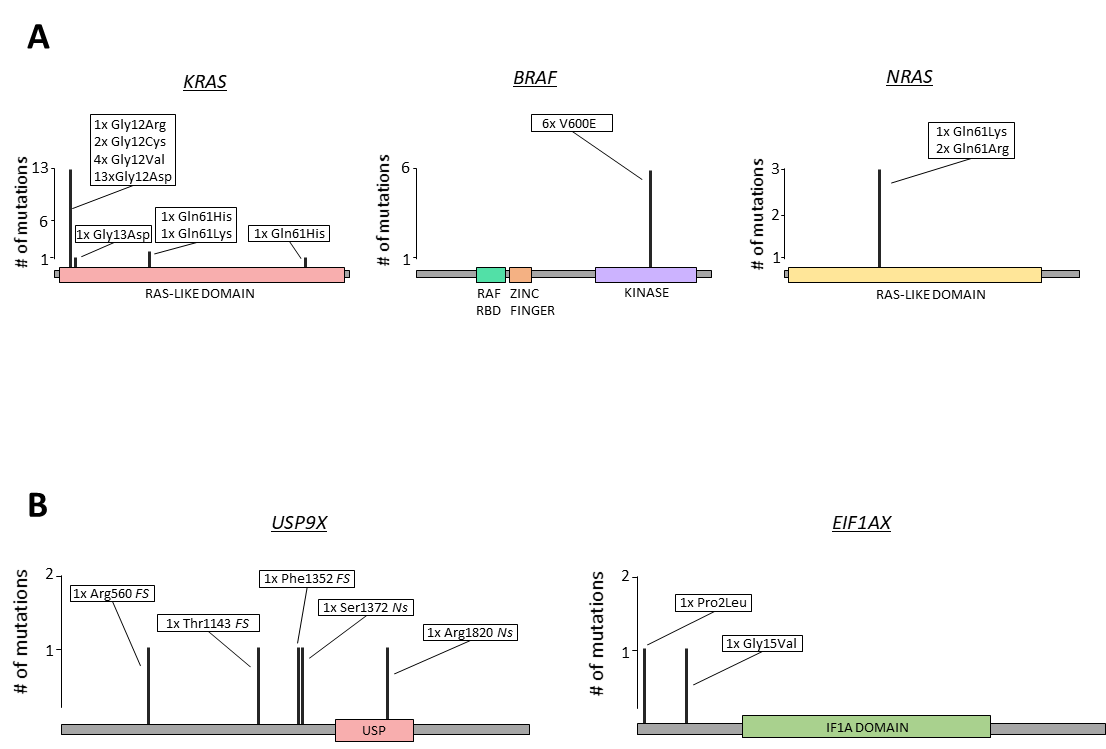

Figure S4. Lollipop plots of mutations across commonly mutated genes. (A) Canonical MAPK pathway genes *KRAS*, *BRAF* and *NRAS*. (B) *USP9X* and *EIF1AX*. Bar height relates to frequency of mutation at each site. Known protein coding domains are labelled throughout genes. Amino acid changes are displayed within boxed areas as reference amino acid, amino acid position, cancer altered amino acid. *FS* = frame shift, *Ns* = nonsense. Note, EIF1AX contains 3 mutations, one of which is a splice site variant which is not shown on the above plot.

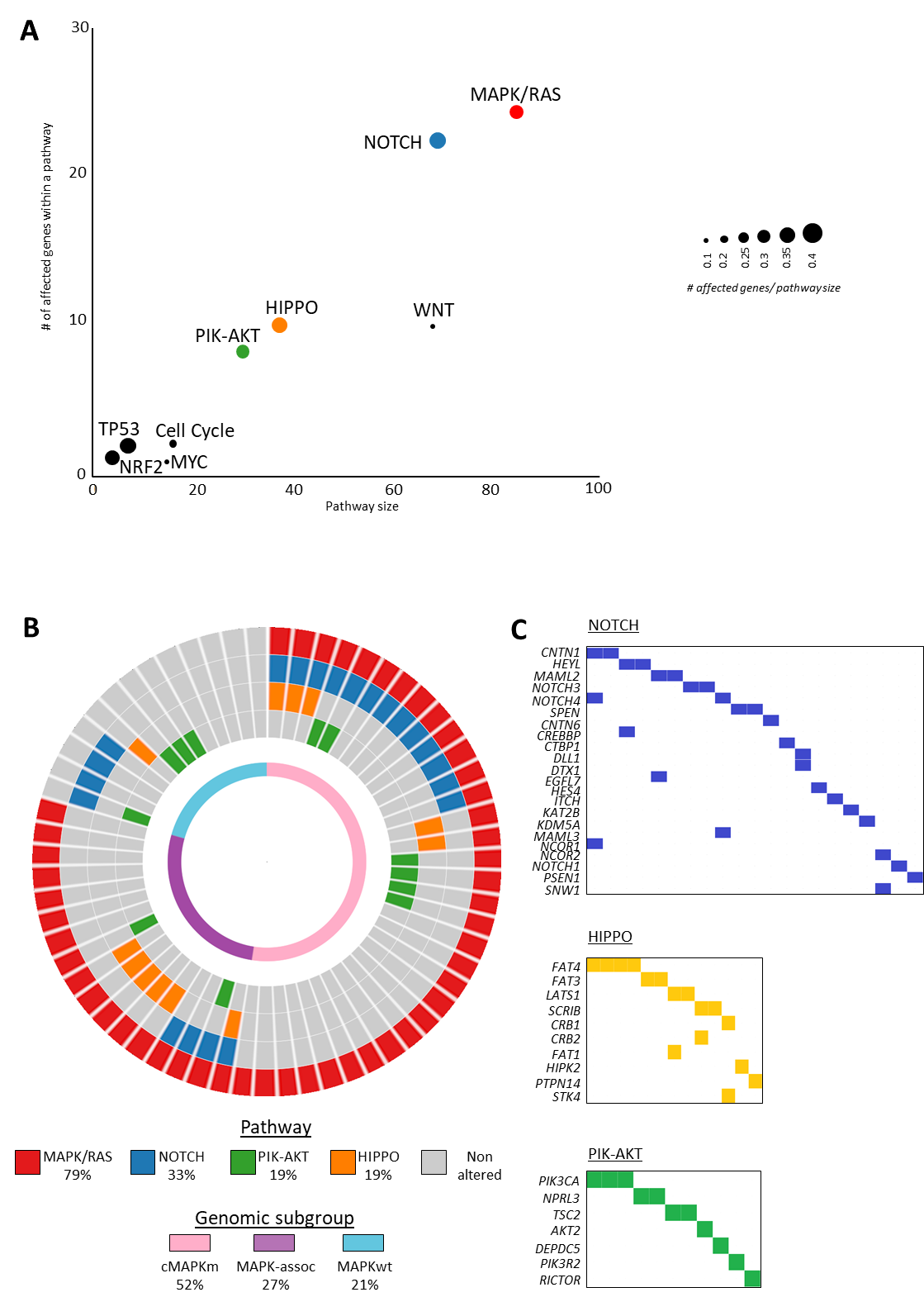

Figure S5. Pathway analysis of genes mutated in low grade serous ovarian carcinoma. A. Scatter plot of the number of genes altered in oncogenic pathways vs total pathway size (size of dot proportional). B. Circular plot of the top four most frequently affected pathways across samples. C. Oncoplot of perturbed genes across the NOTCH, HIPPO and PIK-AKT pathways.

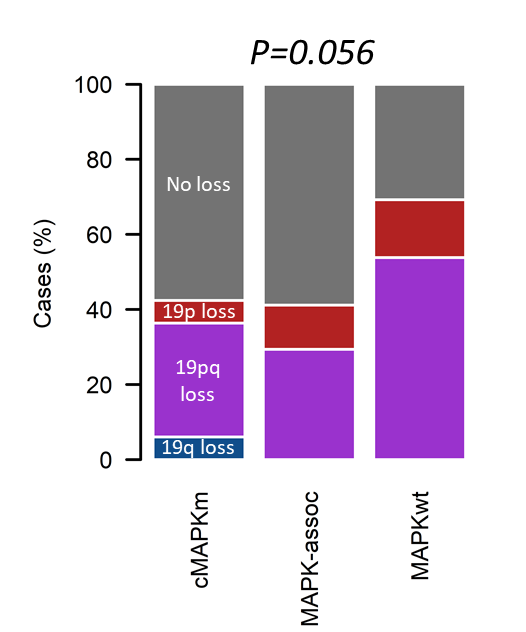

Figure S6. Chromosome 19 aberrations across LGSOC subtypes. Labelled P value represents comparison of chr19p loss (alone or co-loss with chr19q) in cMAPKm vs MAPKwt cases. cMAPKm, canonical MAPK mutant; MAPK-assoc, MAPK-associated mutation; MAPKwt, MAPKwild-type.

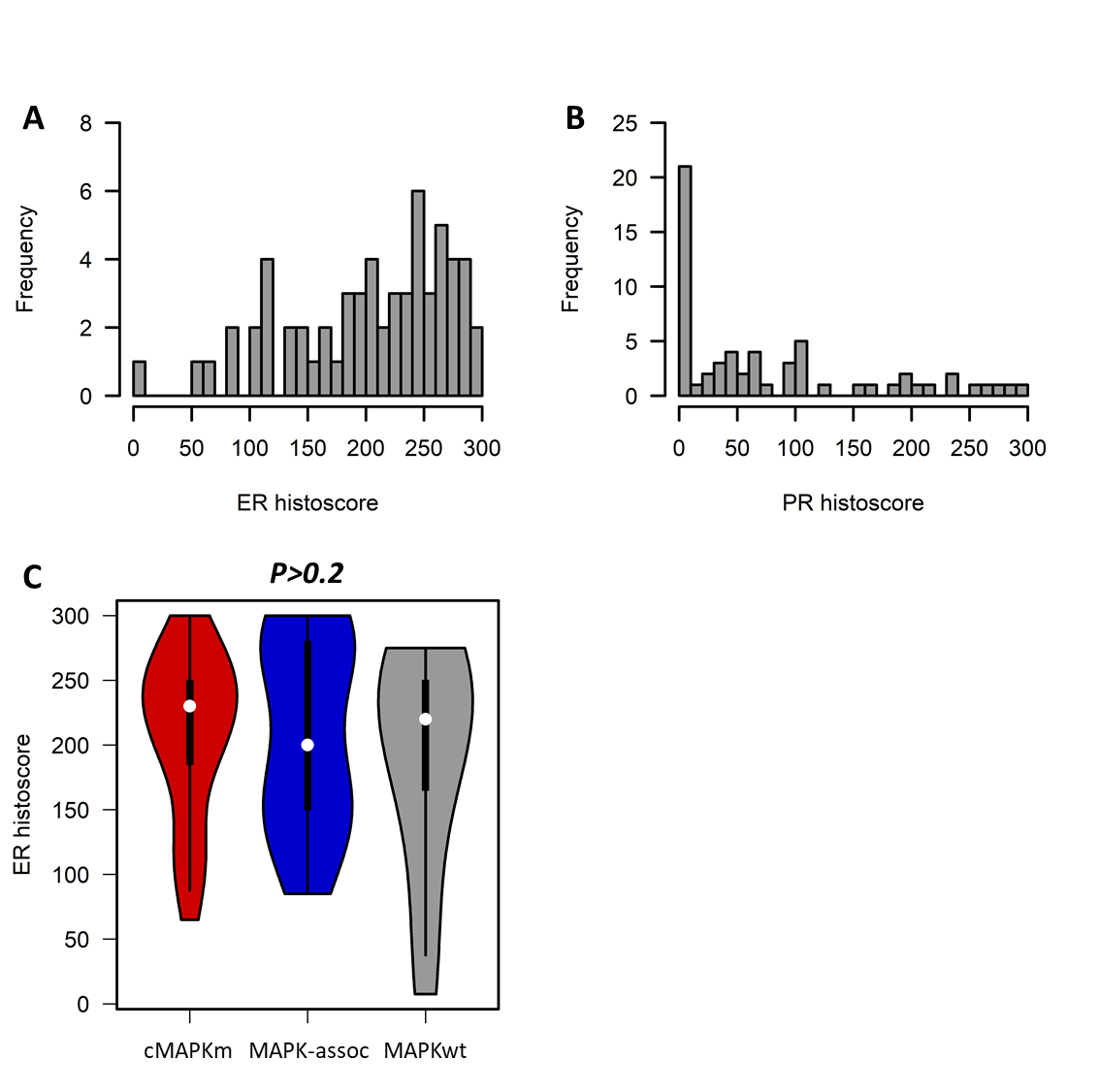

Figure S7. Hormone receptor expression across LGSOC cases. (A) Oestrogen receptor (ER) expression across all cases. (B) Progesterone receptor (PR) expression across all cases. (C) ER expression across genomic subclasses. cMAPKm, canonical MAPK pathway mutation; MAPK-assoc, MAPK-associated mutation; MAPKwt, MAPK pathway wild-type.

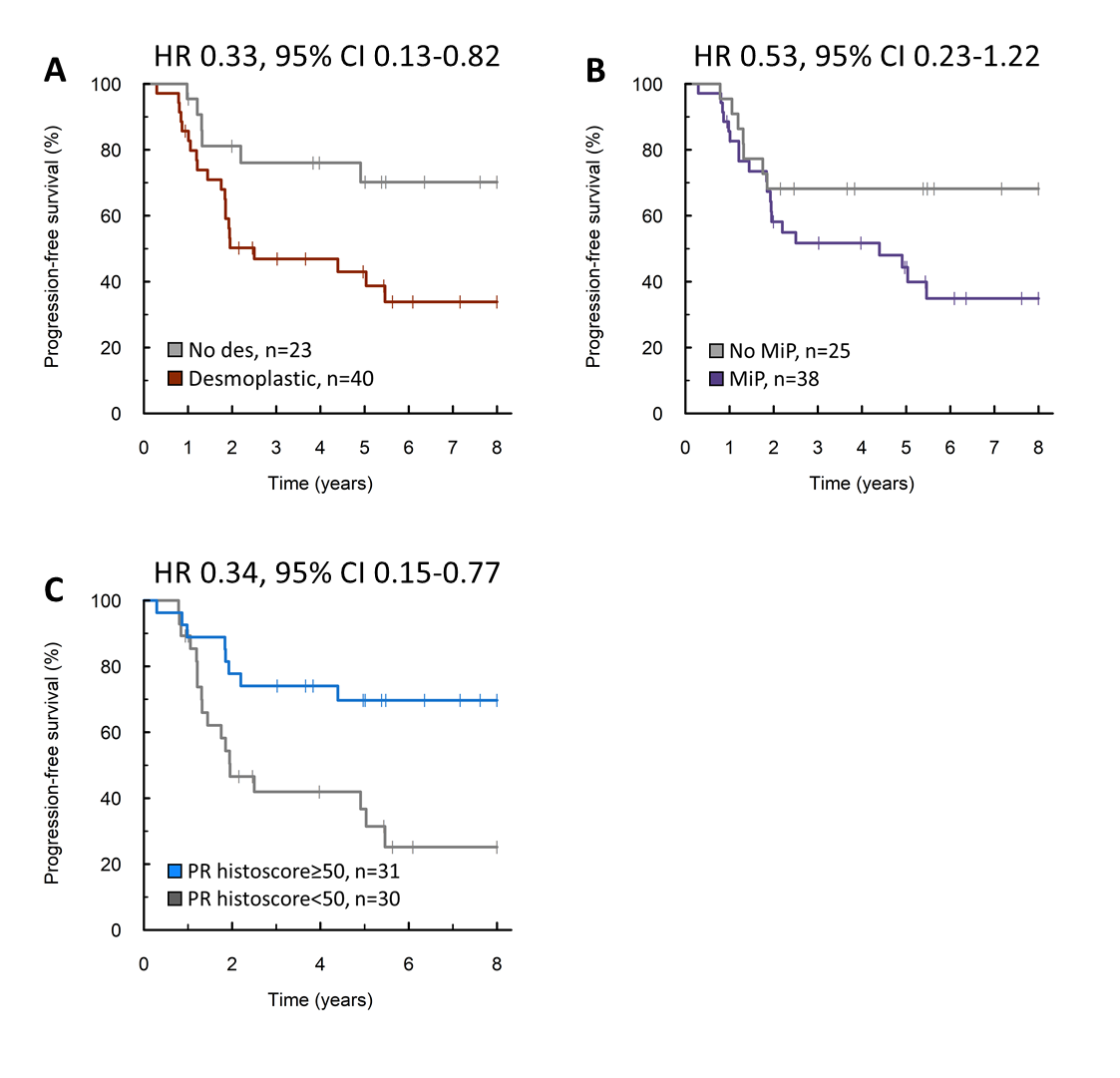

Figure S8. Impact of low grade serous carcinoma features on progression-free survival. (A) Survival time of patients with tumours demonstrating desmoplasia. (B) Survival time of patients with tumours demonstrating the micropapillary (MiP) histological pattern of invasion. (C) Survival time of patients whose tumours express progesterone receptor (PR). HR, hazard ratio; 95% CI, 95% confidence interval.

**Supplementary tables**

Table S1. Clinicopathological features of LGSOC by genomic subtype

|  | | cMAPKm | | MAPK-assoc | | MAPKwt | |
| --- | --- | --- | --- | --- | --- | --- | --- |
|  | | **N** | **%** | **N** | **%** | **N** | **%** |
| Cases | Total | 33 | - | 17 | - | 13 | - |
| Age | Median years | 62 | Range 23-82 | 49 | Range 19-82 | 47 | Range 27-69 |
| FIGO stage | I | 3 | 9.4 | 3 | 17.6 | 3 | 23.1 |
|  | II | 2 | 6.3 | 1 | 5.9 | 2 | 15.4 |
|  | III | 22 | 68.8 | 11 | 64.7 | 6 | 46.2 |
|  | IV | 5 | 15.6 | 2 | 11.8 | 2 | 15.4 |
|  | NA | 1 | - | 0 | - | 0 | - |
| RD following cytoreduction | Macroscopic RD | 19 | 61.3 | 9 | 52.9 | 6 | 46.2 |
|  | No macroscopic RD | 12 | 38.7 | 8 | 47.1 | 7 | 53.8 |
|  | NA | 2 | - | 0 | - | 0 | - |

cMAPKm, canonical MAPK mutant; MAPK-assoc, MAPK-associated mutation; MAPKwt, MAPKwild-type. NA, not available; RD, residual disease

Table S2. Multivariable analysis of disease-specific survival

|  | | HR | 95% confidence interval | P |
| --- | --- | --- | --- | --- |
| Genomic class | cMAPKm | 0.24 | 0.07-0.79 | 0.0195 |
|  | MAPK-assoc | 0.24 | 0.06-0.98 | 0.0467 |
|  | MAPKwt | Ref | Ref | Ref |
| Desmoplasia | Yes | Ref | Ref | Ref |
|  | No | 0.20 | 0.04-0.89 | 0.0343 |
| Micopapillary invasion pattern | Yes | Ref | Ref | Ref |
|  | No | 0.27 | 0.08-0.91 | 0.0355 |
| PR histoscore | ≥50 | 0.38 | 0.12-1.14 | 0.0840 |
|  | <50 | Ref | Ref | Ref |
| FIGO stage | I/II | Ref | Ref | Ref |
|  | III/IV | 0.69 | 0.09-5.41 | 0.7237 |
| RD following cytoreduction | Macroscopic RD | Ref | Ref | Ref |
|  | No macroscopic RD | 0.19 | 0.05-0.79 | 0.0222 |

HR, hazard ratio; FIGO, International Federation of Gynecology and Obstetrics; RD, residual disease
